# A Novel Association Between Human Papillomavirus and Thyroid Eye Disease

**DOI:** 10.1101/2024.04.27.24306443

**Authors:** Ishita Garg, Benjamin I. Meyer, Ryan A. Gallo, Sara T. Wester, Daniel Pelaez

## Abstract

**Context:** Thyroid eye disease (TED) is an autoimmune disease characterized by orbital inflammation and tissue remodeling. TED pathogenesis is poorly understood but is linked to autoantibodies to thyroid-stimulating hormone receptor (TSHR) and insulin-like growth factor 1 receptor (IGF-1R).

**Objective:** To explore the potential involvement of viral infections in TED pathogenesis.

**Methods:** Using NCBI BLAST, we compared human TSHR and IGF-1R proteins to various viral proteomes, including *Papillomaviridae*, *Paramyxoviridae*, *Herpesviridae*, *Enterovirus*, *Polyomaviridae*, and *Rhabdoviridae*. Enzyme-linked immunoassays (ELISAs) were performed on orbital adipose tissue samples from 22 TED patients and controls to quantify antiviral antibody titers. Demographics and clinical data were reviewed.

**Results:** Homology analysis revealed conserved motifs between TSHR and IGF-1R with several viral proteins, particularly the human papillomavirus 18 (HPV18) L1 capsid protein. Basic demographic and clinical information between the cohorts were comparable. ELISAs showed statistically significant differences in the average HPV18 L1 IgG normalized optical density levels among tissues of control (*M* = 0.9387, *SD* = 0.3548), chronic TED (*M* = 2.305, *SD* = 1.064), and active acute TED (*M* = 4.087, *SD* = 2.034) patients. These elevated HPV18 L1 IgG titers did not statistically correlate with TSH, T4, or TSI levels, and were elevated in TED patients irrespective of treatment with teprotumumab, indicating a direct immunological response to HPV.

**Conclusions:** This study presents the first molecular evidence linking HPV and TED, highlighting molecular mimicry between HPV capsid protein and key autoimmunity targets in TED. This suggests an immunological link contributing to TED’s pathogenesis, opening new avenues for understanding and managing the disease.

## Introduction

Thyroid eye disease (TED), also known as thyroid-associated ophthalmopathy or Grave’s orbitopathy, poses a unique clinical challenge for endocrinologists managing systemic thyroid disease. It is characterized by an intricate interplay of autoimmune mechanisms targeting orbital tissues, resulting in debilitating symptoms such as proptosis, diplopia, pain, dry eyes, and redness [1, 2]. In its most severe form, TED can lead to compressive optic neuropathy, potentially causing irreversible vision loss [3]. The incidence of TED in dysthyroid patients varies, ranging from 25% to 50% of individuals with Graves’ disease (GD) and approximately 3% to 5% of those with Hashimoto’s thyroiditis] [4, 5]. TED can also occur in euthyroid and, less commonly, hypothyroid patients [2]. What predisposes an individual to develop TED and what leads to the heterogeneity in clinical manifestations is largely unknown and a subject of significant clinical interest. A better understanding of risk factors for the development of TED in patients with systemic thyroid dysfunction would enable better screening, risk modification, and improved therapeutic management earlier in the disease.

TED is often classified into two distinct clinical phases: an active phase, marked by vasodilation, orbital congestion, and inflammation, and an inactive phase characterized by fibrosis and a potentially reduced responsiveness to therapeutic interventions [6]. The approval of teprotumumab, a monoclonal antibody targeting the Insulin-like Growth Factor-1 Receptor (IGF-1R), by the FDA in 2020 represented a significant advancement in TED management, demonstrating the effectiveness of targeted biological treatments [7]. The phase 2 and phase 3 clinical trials for teprotumumab were conducted with patients with active, inflammatory TED, however recent studies have shown that teprotumumab reduces proptosis in patients with non-inflammatory TED as IGF-1R is overexpressed on fibroblasts in both the inflammatory and non-inflammatory phases [8]. Despite this progress, ongoing clinical trials continue to investigate various pharmacological agents designed to modulate the autoimmune and inflammatory pathways implicated in TED, in pursuit of more comprehensive therapeutic options.

The persistent gaps in our understanding of TED’s pathogenesis, especially regarding the factors influencing its onset and progression, underscore the necessity for further exploration into its underlying mechanisms. Although the majority of TED cases are related to GD, with documented upregulation of IGF-1R and Thyroid-Stimulating Hormone Receptor (TSHR) by autoantibodies, the occurrence of TED in patients without apparent thyroid dysfunction suggests that additional mechanisms beyond direct thyroid autoimmunity may be involved in its development [2, 3].

Emerging insights into the nature of autoimmune diseases have illuminated the potential role of molecular mimicry, a process where external pathogens trigger autoimmune responses by presenting antigens that resemble host tissues [9]. This concept introduces the hypothesis that infections may act as catalysts for autoimmune conditions like TED through a process that is not yet fully understood, suggested by reports of new onset TED following COVID-19 infection [10]. This study seeks to explore the hypothesis that an infectious agent may underlie the pathogenesis of TED, leveraging mechanisms of molecular mimicry which then may initiate or exacerbate the disease process.

## Materials and Methods

### Homology Search

A protein homology analysis was conducted using the National Center for Biotechnology Information (NCBI) Basic Local Alignment Search Tool (BLAST) to compare the human IGF-1R isoform 1 precursor protein (NP_000866.1) and the human TSHR isoform 1 precursor protein (NP_000360.2) with various viral proteomes.

### Participants and Specimen Collection

The University of Miami Institutional Review Board approved this study (IRB 20110692). This study adhered to the tenets of the Declaration of Helsinki. Orbital adipose tissue was collected from the medial fat of upper or lower eyelids in control patients undergoing blepharoplasty or TED patients undergoing orbital decompression surgery at the Bascom Palmer Eye Institute (Miami, FL). Eleven control patients and eleven patients with TED were included in this study.

### Orbital Adipose Sample Processing

Orbital adipose samples were cut into smaller sections and washed with 1X phosphate buffered saline (PBS). Orbital adipose samples weighing between 60 to 110 mg were used for downstream processing. To obtain protein lysates from the tissues, a Branson 450 Digital Sonifier was used at the following settings: a duty cycle of two seconds on/three seconds off for 40 seconds at 40% amplitude. The sample was then centrifuged for ten minutes at 14,000 x g and the supernatant was collected. Samples were stored in -80°C until further use.

### Measurement of HPV18 L1 IgG

A detergent-compatible (DC) protein assay (Catalog # 5000112) was performed to determine protein concentration in the processed samples. Protein concentrations were normalized to the lowest concentration. Based on BLAST results, the Papillomavirus type 18 L1-Capsid Antibody (IgG) enzyme-linked immunosorbent assay (ELISA; Catalog # NBP2-60106-1Kit, RRID: AB_3095886) was performed to measure human papillomavirus 18 L1 capsid protein (HPV18L1) IgG levels by optical density (OD) in control and TED patient samples. Higher OD corresponded to increased light absorption by the sample, reflective of a higher antibody antigen/antibody concentration. The protocol was followed as directed by the manufacturer.

### Clinical Summary

Clinical data were extracted from patient records. Patient demographics and clinical history were reviewed including age, gender, smoking status, thyroid-stimulating hormone (TSH), thyroxine (T4), and thyroid-stimulating immunoglobulin (TSI) levels, and Clinical Activity Score (CAS). The 7-point CAS scale was used.

### Statistical Analysis

To calculate the normalized OD (to account for the variable avidities, i.e. number of binding sites, of different antibodies), the sample OD value was divided by the geometric mean of the control samples as per ELISA manufacturer instructions. Statistical analyses were computed using GraphPad Prism 10.0.2 (La Jolla, California). The one-way ANOVA with the post-hoc Tukey test or unpaired T test were performed to identify differences between the means of the study groups. Pearson correlation analyses with two-tailed p values were performed.

## Results

### Homology Between IGF-1R/TSHR and HPV Major L1 Capsid

BLAST homology analysis identified several homologous short sequences between human IGF-1R and TSHR with various proteomic components within the following viral proteome assemblies: *Papillomaviridae*, *Paramyxoviridae*, *Herpesviridae*, *Enteroviruses*, *Polyomaviridae*, and *Rhabdoviridae*. There were many similarities between human IGF-1R and TSHR with internal and viral matrix proteins, or to proteins encoded by the viral genome which are expressed in the cytoplasm of infected host cells, which deprioritized these proteins as significant antigenic triggers. However, there were homologous alignments between human IGF-1R/TSHR with the major capsid proteins of the human *Papillomaviridae* family. Notably, two motifs from the HPV L1 capsid protein were highly conserved across all human papillomavirus serotypes and human IGF-1R and TSHR sequences: FG*X*V and I*X*E*X*T+NP (**Fig. 1**).

**Figure 1.**
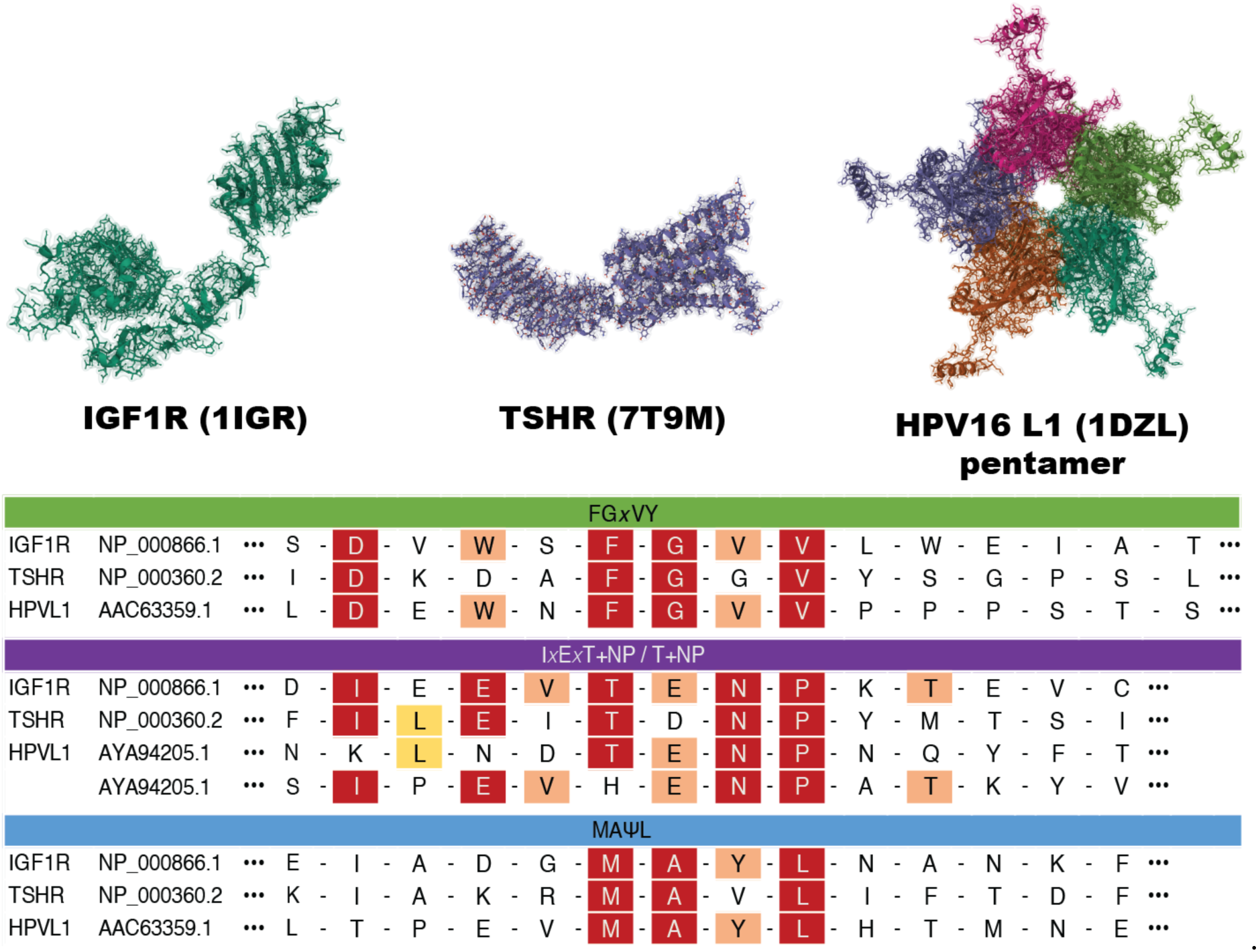
BLAST homology search. Protein homology analysis was conducted using the National Center for Biotechnology Information Basic Local Alignment Search Tool (BLAST) between the human insulin-like growth factor 1 receptor, isoform 1 precursor (accession # NP_000866.1) and the human thyrotropin receptor isoform 1 precursor (accession # NP_000360.2) against the following viral proteome assemblies: *Papillomaviridae*, *Paramyxoviridae*, *Herpesviridae*, *Enteroviruses, Polyomaviridae*, *Rhabdoviridae*. Shown here are the protein motifs (shown in green, purple, and blue) from the HPV L1 capsid protein that are highly conserved across all human papillomavirus serotypes and returned high alignment correlations to motifs in the human IGF-1R and TSHR (*x* = any amino acid; + = similarly charged amino acid; ψ = hydrophobic amino acid).

### Clinical Summary

The demographic and clinical characteristics of the patient cohort are outlined in **Table 1**. 22 patients were included in this study (11 controls and 11 with TED). Acute disease activity was defined as CAS ≥ 4 for less than 12 months; chronic disease was defined as lasting more than 12 months. Inactive disease was defined by patients with a CAS < 3. Of the patients with TED, 7 individuals had chronic disease and 4 individuals had acute active disease. Most patients were female: 10 (10/11; 90.9%) in the control, 6 (6/7; 85.7%) in the chronic TED, and 3 (3/4; 75%) in the acute active TED groups. Mean ages at the time of surgery were 63.02 years (range: 54-70.6), 52.66 years (range: 37.4-74.4), and 63.73 years (range: 49.4-69.8) for control, chronic TED, and acute active TED patients, respectively. When comparing chronic versus acute active TED patients, no statistically significant differences emerged in TSH (p = 0.6862), T4 (p = 0.742), and TSI levels (p = 0.4931). The average highest CAS score for chronic TED patients was 3 ± 2.236 while the average highest CAS score for acute active TED patients was 5.25 ± 0.9574.

**Table 1:**
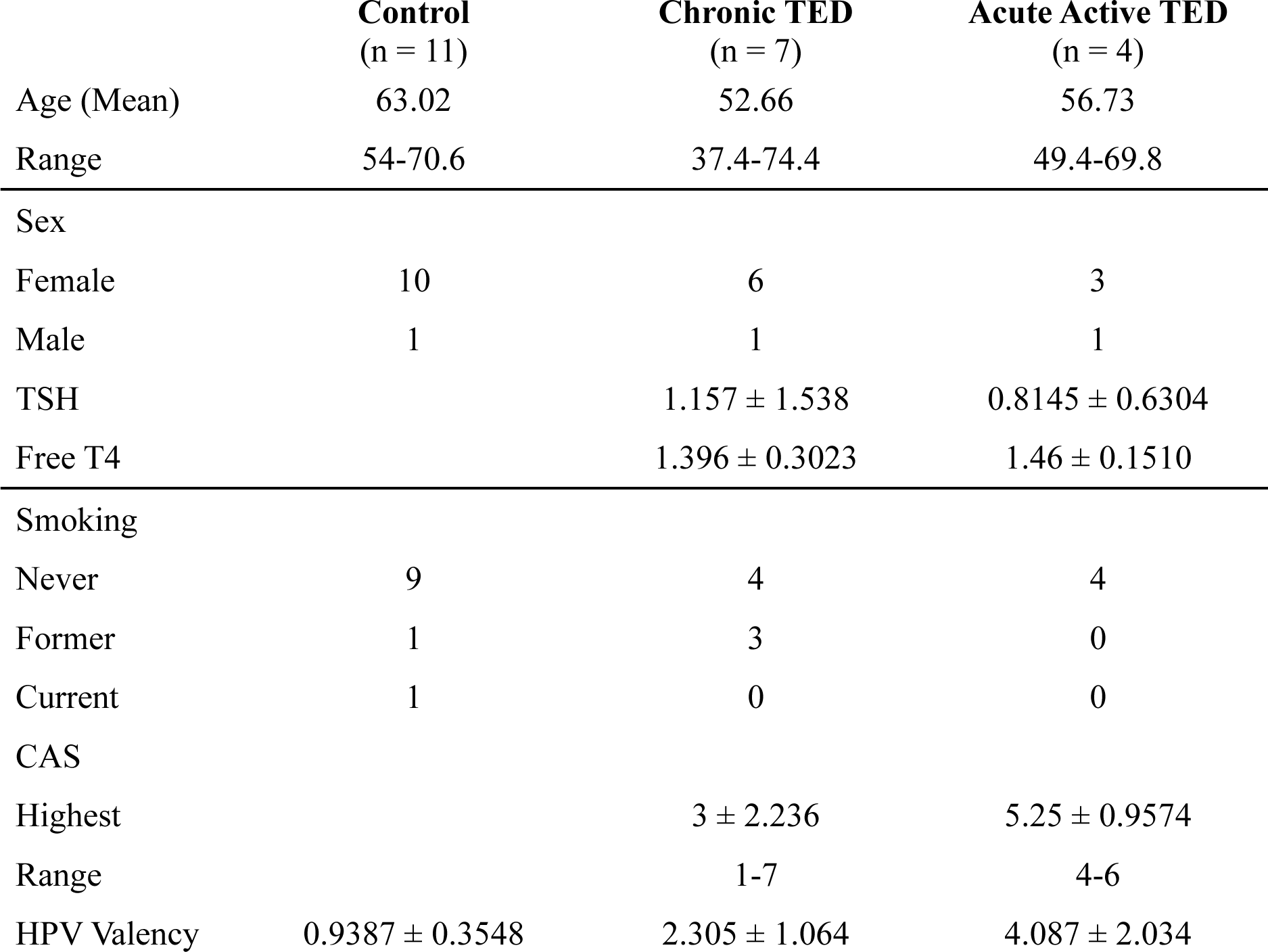
Patient Characteristics.

|  | **Control**<br>(n = 11) | **Chronic TED**<br>(n = 7) | **Acute Active TED**<br>(n = 4) |
| --- | --- | --- | --- |
| Age (Mean) | 63.02 | 52.66 | 56.73 |
| Range | 54-70.6 | 37.4-74.4 | 49.4-69.8 |
| Sex |  |  |  |
| Female | 10 | 6 | 3 |
| Male | 1 | 1 | 1 |
| TSH |  | 1.157 ± 1.538 | 0.8145 ± 0.6304 |
| Free T4 |  | 1.396 ± 0.3023 | 1.46 ± 0.1510 |
| Smoking |  |  |  |
| Never | 9 | 4 | 4 |
| Former | 1 | 3 | 0 |
| Current | 1 | 0 | 0 |
| CAS |  |  |  |
| Highest |  | 3 ± 2.236 | 5.25 ± 0.9574 |
| Range |  | 1-7 | 4-6 |
| HPV Valency | 0.9387 ± 0.3548 | 2.305 ± 1.064 | 4.087 ± 2.034 |

### Increased anti-HPV18 L1 IgG Titers in Chronic and Active TED Patients

Based on the results from the BLAST homology search, we next measured IgG antibody titers against HPV18 L1 capsid protein by ELISA in orbital adipose tissue from the control and TED patients. The mean normalized optical densities of human HPV18 L1 IgG antibody in the orbital adipose tissues were 0.9387 ± 0.3548, 2.305 ± 1.064, and 4.087 ± 2.034 from the control, chronic TED, and acute active TED groups, respectively. These mean concentrations were significantly different between control and chronic TED patients (p = 0.0344), control and active acute TED patients (p = 0.0001), and chronic and active acute TED patients (p = 0.0333) (**Fig. 2a**). There was no significant evidence for correlations between orbital adipose tissue HPV18L1 IgG valency with TSH levels, T4 levels, or TSI levels in either chronic or acute active TED patients (**Fig. 2b**; **Table 2**).

**Figure 2a:**
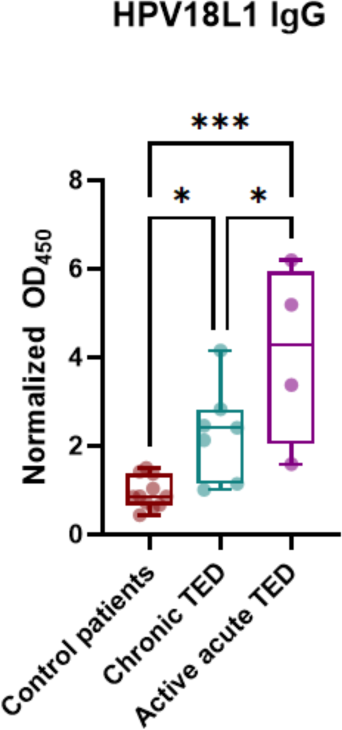
Human papillomavirus 18 L1 capsid protein (HPV18L1) IgG levels orbital fat samples. P value denoted by: * is ≤ 0.05; ** is ≤ 0.01; *** is ≤ 0.001.

**Figure 2b:**
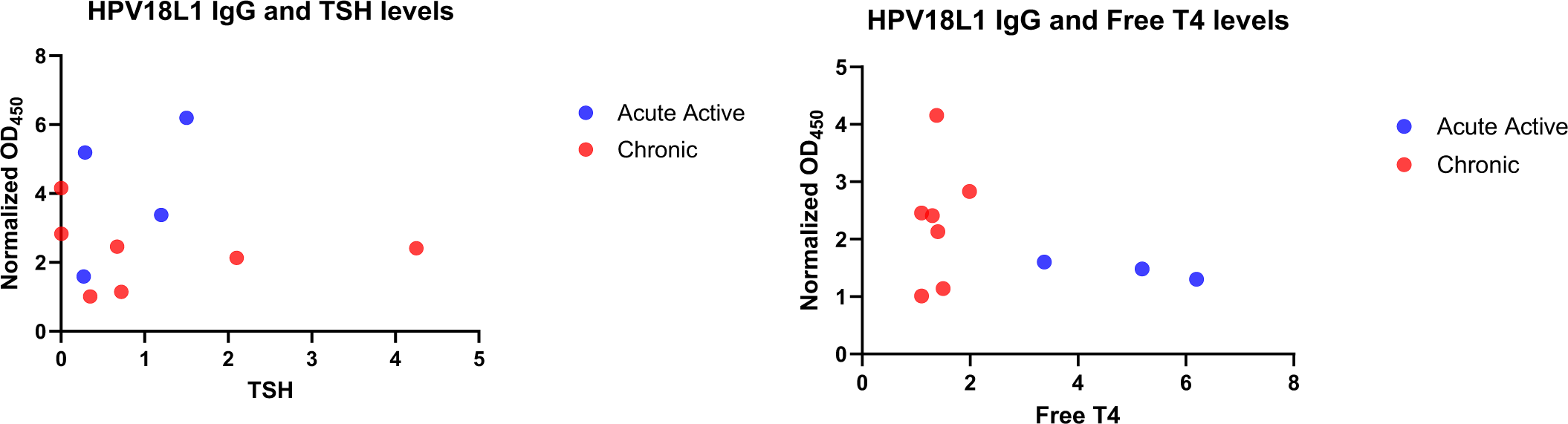
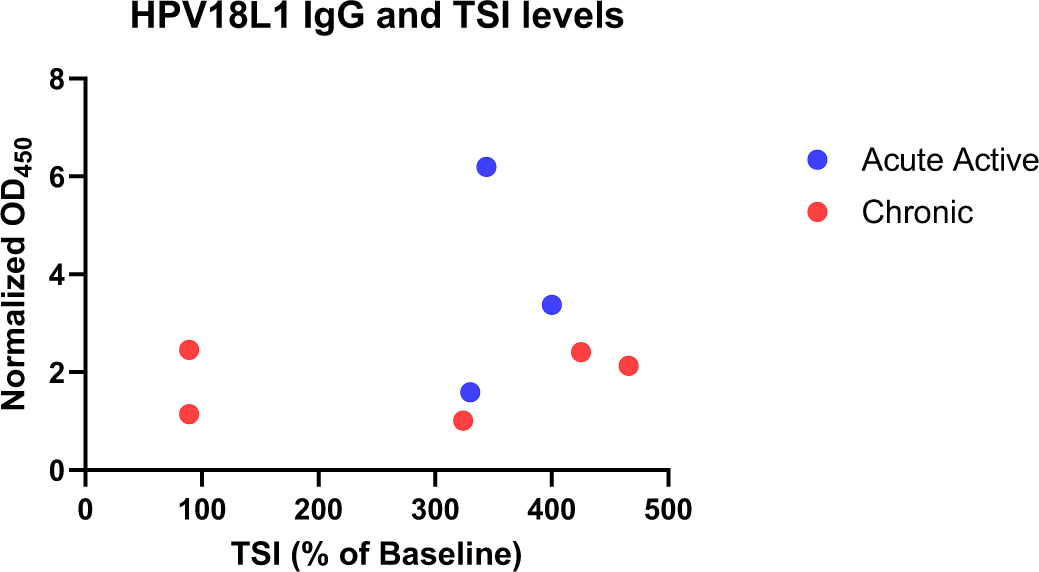
Subgroup analysis comparing thyroid status with HPV18L1 IgG levels. TSH = thyroid stimulating hormone. Normalized OD_450_ is the normalized optical density absorbability as the metric for antibody titer based on the enzyme-linked immunosorbent assay (ELISA)

**Table 2:** HPV18L1 IgG Titer Subgroup Analysis.

|  | **Chronic TED**<br>(n = 7) |  | **Acute Active TED**<br>(n = 4) |  |
| --- | --- | --- | --- | --- |
|  | Correlation<br>coefficient | P value | Correlation<br>coefficient | P value |
| TSH level | -0.5000 | 0.2667 | 0.8000 | 0.3333 |
| T4 level | 0.2536 | 0.5832 | -0.9617 | 0.1768 |
| TSI level | -0.6725 | 0.2136 | -0.1543 | 0.9014 |

To parse out potential effects of teprotumumab intervention in TED patients on IgG HPV18 L1 valency, we performed subgroup analysis (**Fig. 2c**). No significant difference was noted comparing treated versus non-treated patients (Student T test). Further analyses did not reveal statistically significant differences between antibody titers in patients when comparing between radioactive iodine treatment or prior thyroidectomy (**Supplemental Fig 2**).

**Figure 2c:**
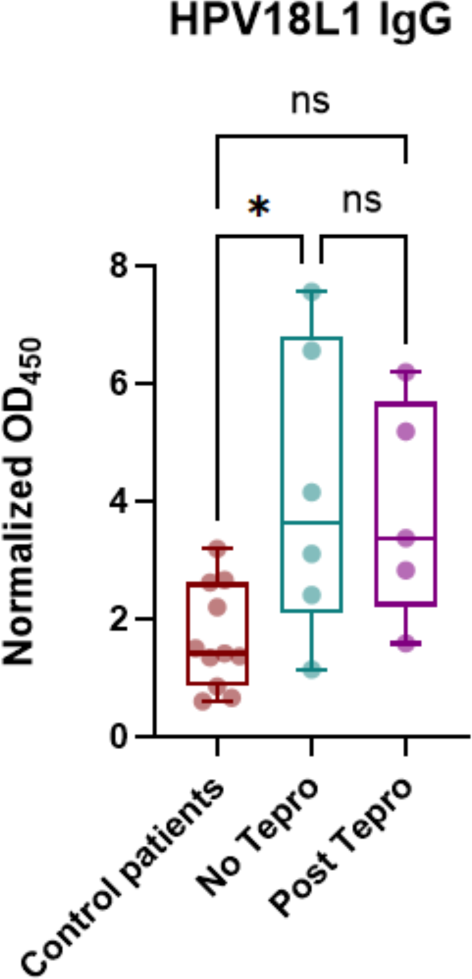
Impact of teprotumumab treatment on HPV18L1 IgG levels. Tepro = teprotumumab. P value denoted by: * is ≤ 0.05; ns: not significant.

## Discussion

This study presents molecular evidence suggesting a novel association between HPV and TED, facilitated potentially through molecular mimicry between the HPV’s L1 capsid protein and significant autoimmunity targets in TED, specifically IGF-1R and TSHR. The discovery of significantly elevated IgG antibodies against the HPV L1 capsid protein in patients with chronic and active TED—irrespective of treatment with teprotumumab—highlights a potential immunological connection that may play a crucial role in TED’s pathogenesis.

The *Papillomaviridae* family encompasses over 150 HPV subtypes, with types 16 and 18 being primary culprits in cervical cancers [11]. Molecular mimicry has been implicated in various autoimmune conditions, including rheumatoid arthritis, systemic lupus erythematosus (SLE), Sjogren’s syndrome, autoimmune liver disease, inflammatory bowel disease, and multiple sclerosis [12]. This phenomenon has also been proposed to explain the rare occurrences of autoimmune disease activation or flares following vaccination; however, such data are scarce and sometimes conflicting, necessitating cautious interpretation to avoid potential confounders or the mistaken extrapolation of correlation for causation [13-16].

The question of molecular mimicry’s role in TED through HPV infection introduces a new dimension to this discussion. Research indicates that individuals with SLE not only show higher rates of high-risk HPV infections but also an increased risk of cervical dysplasia, likely due to immunosuppression [17, 18]. This suggests that an underlying autoimmune or inflammatory state may elevate HPV susceptibility. Moreover, a retrospective study of 62 patients highlighted a significant association between HPV infection and autoimmune disorders, with a 20% prevalence of autoimmune disease in HPV DNA-positive women versus approximately 3% in the general population, alongside increased autoantibody levels, underscoring the complex link between HPV and autoimmunity [19].

The widespread vaccination against high-risk HPV strains, aimed at preventing cervical cancer, has yielded substantial data for analysis, particularly in young women who are more susceptible to autoimmune diseases due to hormonal changes during puberty [20]. Currently, three HPV vaccines are in use: Cervarix (bivalent), Gardasil (quadrivalent), and Gardasil9 (nonavalent) [11]. A significant study by Jacobsen et al., involving nearly 190,000 vaccinated young women, reported six new cases of Graves’ disease post-vaccination with the quadrivalent vaccine [21]. However, in a separate study this group found no pattern of autoimmune diseases related to vaccine timing, dosage, or patient age, suggesting that the observed autoimmune thyroid cases might have pre-existed vaccination, as indicated by a non-elevated incidence rate ratio (IRR=0.72 [0.50-1.01]) upon detailed review [22].

The quadrivalent HPV vaccine, which leverages virus-like particles from the HPV L1 capsid protein, aims to replicate the HPV virus’s structure to stimulate a suitable immune response [23, 24]. Our findings show a homology between the HPV L1 capsid protein and key TED autoimmunity targets (IGF-1R and TSHR), suggesting potential autoimmune disease triggers from HPV L1 protein exposure or vaccination. Yet, prior extensive analyses, including a systematic review and meta-analysis of over 169,000 autoimmune disorder cases, indicated no significant risk increase for autoimmune diseases post-HPV vaccination (odds ratio 1.003) [25]. This suggests HPV vaccination is safe and unlikely to directly initiate autoimmune disease, though it may reveal pre-existing autoimmune conditions, necessitating further research. These insights highlight the need to differentiate the effects of natural HPV infection from vaccination outcomes in TED research, ensuring that vaccine-induced autoimmunity is not mistakenly implicated or deduced.

The higher prevalence of both HPV and TED in females, contrasted with more severe disease manifestations in males, raises intriguing questions about sex-specific immune responses and hormonal influences on disease severity and progression [26]. This sex disparity could provide critical insights into the underlying mechanisms of HPV’s impact on TED, highlighting an area ripe for further investigation.

Our study faces several limitations, including a small sample size and an imbalance in male-to-female ratios (albeit reflective of natural demographic differences), which may affect the generalizability of our findings. Additionally, the absence of detailed clinical histories regarding prior HPV infection or vaccination status among participants limits our ability to fully understand the relationship between HPV exposure and TED development.

Despite these limitations, our findings offer a groundbreaking perspective on TED’s association with HPV through molecular mimicry, establishing a foundation for subsequent, more definitive research. This exploratory study serves as a catalyst for future investigations to confirm the causative link between HPV and TED, potentially revolutionizing our approach in managing this complex autoimmune condition.

## Data Availability

All datasets generated and analyzed during the current study are not publicly available but are available from the corresponding author on reasonable request.

## Acknowledgements/Funding

This work was supported in part by the Dr. Nasser Al-Rashid Orbital Research Endowment (Miami, FL, USA). Bascom Palmer Eye Institute is supported by NIH Center Core Grant P30EY014801 and a Research to Prevent Blindness Unrestricted Grant (New York, NY, USA).

## Supplemental Figures

**Supplemental Figure 1.**
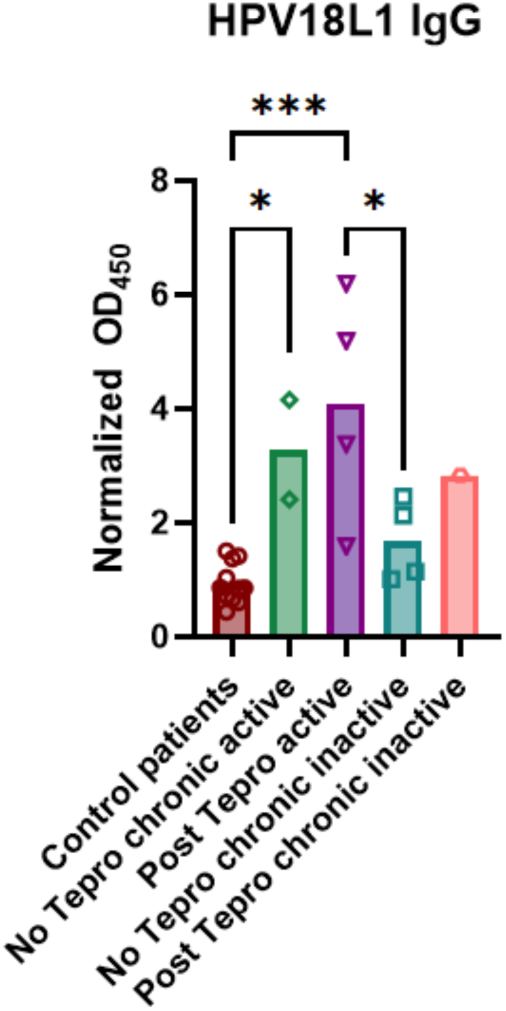
Subgroup analysis of teprotumumab treatment and thyroid eye disease status with HPV18L1 IgG levels.

**Supplemental Figure 2.**
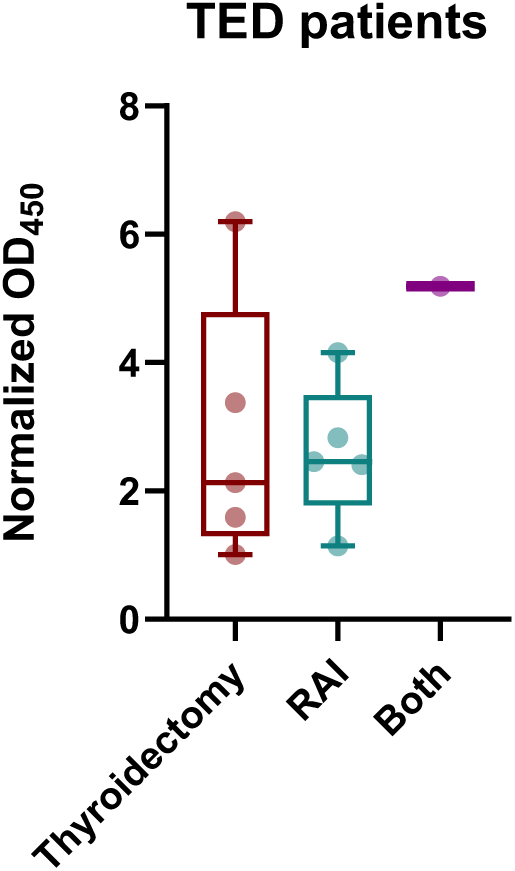
Impact of thyroid treatment on HPV18L1 IgG levels.

## Notes

### Competing Interest Statement

The authors have declared no competing interest.

### Author Declarations

IRB of the University of Miami gave ethical approval for this work.

